# New models of transmission of COVID-19 with time under the influence of meteorological determinants

**DOI:** 10.1101/2020.05.26.20113985

**Authors:** Atin Adhikari, Shilpi Ghosh, Moon M. Sen, Rathin Adhikari

## Abstract

**Objectives:** This work aimed at modeling the progressions of COVID-19 cases in time in relation to meteorological factors in large cities of Brazil, Italy, Spain, and USA, and finding the viability of SARS-CoV-2 virus in different weather conditions based on models.

**Methods:** New models constructed showing the relationship of the *I*′ (the number of infected individuals divided by the total population of a city) with the independent variables -time, temperature, relative humidity, and wind velocity. The regression models fitting in the data were statistically validated by: 1) plot of observed and predicted response; 2) standardized residual plots showing the characteristics of errors; 3) adjusted 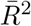 value; 4) the *p* value for the parameters associated with the various independent variables; and 5) the predictive power of the model beyond data points.

**Results:** Models indicate that 1) the transmission of COVID-19 could be relatively high either for elevated temperatures with lower relative humidity or for lower temperatures with higher relative humidity conditions; 2) disease transmission is expected to be reduced more with higher wind velocity; 3) the rate of increase in the number of COVID-19 cases increases in one model with a constant rate and in the other two with varying rates in time. These transmission features seem to have connections with the structural components of the SARS-CoV-2 virus. Under suitable meteorological conditions, the partial natural disappearance of COVID-19 pandemic could be possible.

**Conclusion:** New models for *I′* may be considered to understand the viability of the virus in the environment and future transmission of COVID-19.

## 1. Introduction

The highly pathogenic coronavirus disease 2019 (COVID-19) has become a pandemic after its initial outbreak in Wuhan, Hubei province of China, during December, 2019. According to the recent report of the World Health Organization, the disease has spread to six continents and 210 countries and as on 10 August 2020, there have been 19,687,156 confirmed cases of COVID-19, including 727,435 deaths (WHO, 2020). The causative organism for COVID-19 is severe acute respiratory syndrome coronavirus 2 (SARS-CoV-2), a genus belonging to family Coronaviridae. Clinically, patients with COVID-19 develop respiratory symptoms, which is very similar to other respiratory virus infections. Multiple symptoms may be involved with COVID-19, including respiratory (cough, shortness of breath, sore throat, rhinorrhea, hemoptysis, and chest pain), gastrointestinal (diarrhea, nausea, and vomiting), musculoskeletal (muscle ache), and neurologic (headache or confusion) types. More common signs and symptoms are fever (83% - 98%), cough (76% - 82%), and shortness of breath (31% - 55%) (Wu et al. 2020). Historically, coronaviruses gained prominence during the outbreak of severe acute respiratory syndrome (SARS) during 2002-2003 with severe acute respiratory syndrome coronavirus (SARS-CoV) as the causative agent. The virus infected 8098 individuals with a mortality rate of 9% across twenty-six countries worldwide. In contrast, the incidence of COVID-19 infection has crossed more than 4.42 million to date worldwide, indicating increased transmission ability of SARS-CoV-2 (Cheng et al.,2007; Walls et al., 2020).

According to recent evidence, SARS-CoV-2 virus is primarily transmitted between people through respiratory droplet, direct contact with infected people and indirect contact with surfaces in the immediate environment or with objects used on the infected person (Chan et al., 2020; Li et al., 2020; Liu et al., 2020). Some scientific studies have provided the initial evidence for the viability of SARS-CoV-2 virus in aerosols for hours, suggesting their plausible airborne transmission (van Doremalen et al., 2020). Besides, their human transmissibility can be influenced by the environment in which pathogen and host meet (Pica and Bauvier 2012). Like other influenza virus infections, relative humidity and temperature are expected to affect the incidence of COVID-19 particularly through airborne respiratory droplets (Casanova et al., 2010; Pica and Bauvier, 2012; Lowen and Steel, 2014). The temperature and relative humidity could affect the physicochemical characteristics of infectious droplet including, pH and salt concentration of droplets as well as viral membrane lipid and surface proteins and thus influencing their transmission (Lowen and Steel, 2014; Yang and Marr, 2012). Apart from temperature and relative humidity, the wind velocity also could play a role in the transmission of the virus.

In most of the earlier works related to the spreading of the epidemic, the rate of increase of the number of infected individuals has been studied in relation to the various meteorological determinants (Wang et al., 2020; Prata et al., 2020; Sajadi et al., 2020). However, the rate would vary between cities because of their differences of total population, area and population density. Hence, in the present communication a different approach has been followed to study the spreading of COVID-19 by considering homogeneous mixing in the populations of various major cities (Spain, Italy and the USA) along with the meteorological determinants and population density. Under this consideration, the relevant quantity is the proportionate mixing of the number of infected individuals *I*, within the total population *N* of a city, denoted as *I′* which is equal to 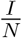. In this work, in the context of infection due to SARS-CoV-2 virus, we have found the relationship between *I′* with temperature, relative humidity, wind velocity and time.

COVID-19 cases in the major cities of four countries have been considered for the statistical analysis and building models. The major cities have higher number of COVID-19 cases and with respect to large population size, almost homogeneous mixing of the population may be expected. On the other hand, small cities have different heterogeneous issues of the population, like change in population density, social behavior and movement of people could be more manifested, which are complicated to be taken into account for the statistical analysis. As our primary concern is to find the response of the SARS-CoV-2 to different meteorological factors as well as the global properties of the spreading of the virus over space and time, the major cities with a higher population are expected to be more appropriate for the statistical analysis of the data on number of infected cases.

In section 2, the data sources and their collection time period are mentioned. In section 3, regression models are built up from the statistical analysis of the data on the number of infected persons in various cities, the related meteorological data and the time elapsed after the initial reporting of infected cases. Mainly based on standardized residual plots and *p* values of numerical coefficients of different independent variables, we have found out the most suitable model in which the proportionate mixing *I*′ could depend on temperature, relative humidity, wind velocity, and time in a statistically significant way. In section 4, the results of the statistical analysis corresponding to various models have been presented with the help of different contour plots, which show the combination of temperature, relative humidity and wind velocity for favourable or unfavourable COVID-19 transmission. Also it has been shown that how the number of infected cases for a particular population, could evolve with time under various meteorological conditions. In section 5, it has been discussed through illustrative example, how models indicate the possibility of the partial natural disappearance of COVID-19 due to the change in the meteorological factors in the environment. In section 6, the connections of transmission characteristics and viability of virus under different meteorological conditions, with the structural components of the SARSCoV-2 virus, are discussed. Finally, in section 7, based on models, concluding remarks with some precautionary measures have been mentioned.

## 2. Data sources and time period of data collection

All data used for analysis were available in public databases. The data on the number of COVID-19 infection cases of several major cities in Spain, Italy, and the USA (Countries with relatively severe COVID-19 outbreak) from the following sources -(1) Data of Spain (URL: https://www.mscbs.gob.es/profesionales/saludPublica/ccayes/alertasActual/nCov-China/documentos/Actualizacion_122_COVID-19.pdf) were collected from the database of the Centro de Coordinací on de Alertas y Emergencias Sanitarias (CCAES), Spain. The center is responsible for coordinating information management and supporting the response to national or international health alert or emergencies, (2) Data of Italy (URL: http://www.salute.gov.it/portale/nuovocoronavirus/dettaglioContenutiNuovoCoronavirus.jsp?area=nuovoCoronavirus&id=5351&lingua=italiano&menu=vuoto) were collected from the database of The Ministry of Health (Italian: Ministero della Salute), which is a governmental agency of Italy and is led by the Italian Minister of Health. (3) Data on different cities of USA (URL: https://usafacts.org/visualizations/coronavirus-covid-19-spread-map/) were collected from USAFacts, which is a not-for-profit, nonpartisan civic initiative providing the most comprehensive and understandable source of government data available in the US. Data on cumulative COVID-19 positive cases, population, and population density for Sau Paulo, Rio de Janeiro, and Brasilia, Brazil, were collected from Wikipedia (https://en.wikipedia.org/wiki/COVID-19_pandemic_in_Brazil/Statistics). Meteorological data for all above-mentioned locations were collected from the World Weather Online database (URL: www.worldweatheronline.com).

We have considered data of the cities Madrid, Catalonia and Pais Vasco-Basque in Spain during the period 4th March, 2020 to 14th March, 2020 before lockdown/travel restrictions and 15th March to 29th March after lockdown/travel restrictions. We have considered data of the cities Milan, Bologna and Venice in Italy during the period 26th February, 2020 to 9th March, 2020 before lockdown and 10th March to 31st March after lockdown. We have considered data of the cities of New York, San Francisco, Atlanta, Seattle, Chicago and Los Angeles in the USA during the period 2nd March, 2020 to 14th March, 2020 before travel restrictions/stay at home order orders. Data of the cities of New York, Chicago, Los Angeles in USA were also considered during the period 16th March to 31st March after travel restrictions/stay at home orders. We have considered data of the cities of Brazil as follows: Sau Paulo during the period 26th February, 2020 to 23rd March, 2020, Rio de janeiro during the period 5th March, 2020 to 1st April, 2020 and Brasillia during the period 7th March, 2020 to 5th April, 2020.

## 3. Regression Models for *I*′

Although we have considered data of different cities both before and after lockdown/travel restrictions, the global features of the spreading of COVID-19 in relation to climatic conditions should be examined before the implementation of restrictions when there were no human interventions on the transmission of the virus. However, assuming that the early parts of restrictions were not so stringent in the selected large cities, the combination of both before and after restrictions data have been considered for statistical analysis. But while interpreting any results from the analyses of the combined data, one needs to be careful about this assumption.

In building up regression models, our approach is different from the conventional models like *SIR* models (Anderson and May,1979; Jong et al., 1995; Dietz 1985). *S*, *I* and *R* correspond to the number of susceptible, infected and recovered individuals, respectively. In some modified models, the number of deaths due to the viral infection has also been considered. As the aim of this work is to determine the influence of the meteorological factors on the spreading of virus infection, we are mainly concerned with the number of infected individuals *I*. The models of transmission are related to the number of infected cases at early stage when the role of virus is highly important. At early stage of transmission of virus, the number of recovered persons or the number related to mortality are much smaller than the number of infected individuals and also these numbers are more related to health of the infected persons, health facilities etc., and are not related to the virus explicitly. Although these numbers do influence *S* in the early period of the pandemic (which we are considering), they can be easily ignored because of being relatively much smaller than *I*. In finding the connection of the spreading of the virus with the meteorological factors and time, the proportionate mixing *I*′ = *I*/*N* will be considered as a parameter for the spreading of virus and, instead of I, the evolution of the parameter *I*′ will be studied. This has the advantage as the upper limit of the quantity *I*′ is normalized to unity for a total population *N* in any city. It is legitimate to assume that the evolution of *I*′ in different cities will follow almost same dynamics with respect to the variation of meteorological factors and time. So same regression model for *I*′ is expected to be valid for different cities and in building such model, different data of different major cities on the number of infected cases may be considered all together for statistical analysis.

We have not considered a priori any specific form for evolution of *I*′ with time *Ti*, like conventional model that takes into account an exponential increase in the number of infected cases with respect to time. The reason behind this is that in the SIR models, the rate of increase in the number of infected persons *I* with time *Ti* will be proportional to the proportion of infectious contacts (*I/N*). The model assumes that further infections occur due to the direct contact with the infected people and one follows the principle of mass action in chemistry where the proportionate mixing *I/N* stands for the concentration of some substance. But the spread of viral infections is due to the contacts of a large number of viruses with the population *N* and may be compared to some extent, with the scattering of large number of elementary particles. This is because infections will be happening due to virus which could be in the air, which could be on the surface of some material, which could be due to the large number of viruses coming from the nasal and respiratory droplets. Furthermore, viruses from one infected person, may infect a large or a few number of people depending on the contacts with the numbers of individuals. The rate of increase in the number of infected individuals should not be considered to be proportional to the number of already infected persons *I*. So considering the viral infection problem as the one similar to the law of mass action in chemistry may not be correct and the rate of increase may not be proportional to *I/N*. In that case, the exponential time dependence which is a solution to *I* coming from the first order differential equation in *I* and time *Ti* in which 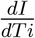 is proportional to *I*, may not be correct. For these reasons, in building up regression models, in the time evolution part of *I*′, we have explored other possible functional forms with time *Ti* as variable. However, we have also explored the possibility of exponential time dependence, although that is not necessarily equivalent to considering that 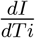 is proportional to *I* and will be explained further in the later part of the paper.

In the absence of our understanding of the relationship of *I*′ with time *Ti* at this point of discussion, *I*′ is considered to be depending on two different functions of time: *F*_1_(*Ti*) and *F*_3_(*Ti*). Also *I*′ is expected to depend on various meteorological factors like temperature (*T* in ^0^*C*), relative humidity (*H* in percentage) and wind velocity (*W* in Km/hr) and for that another function *F*_2_(*T, H, W*) is considered which has no explicit *Ti* dependence and are separate from *F*_1_(*Ti*) and *F*_3_(*Ti*). The metorological factors *T, H* and *W* may or may not vary with time and but have no explicit time dependence. In terms of these functions *I*′ is written as

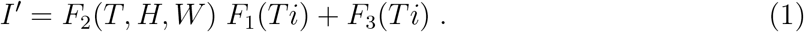

These functions could be related to the interactions of virus with individuals and with environment, viral replication processes and so on. The reason for considering such dependence is as follows. All the terms in the right hand side, should be proportional to *Ti*. This is because at *Ti* = 0, *I*′ is assumed to be zero or very small. In the product of the two functions *F*_1_ and *F*_2_, it is expected that each additive terms should have *Ti* or its higher power as a factor, but *T, H* and *W* may or may not be present. *F*_3_(*Ti*) is another time dependent but independent of meteorological variables. Also because of above reason, both *F*_1_(*Ti*) and *F*_3_(*Ti*) are restricted in the sense that they almost vanish when *Ti* = 0. The rate *R* of increase of *I*′ with time can be written, as the total derivative for which *R* = *R_tot_* is written as

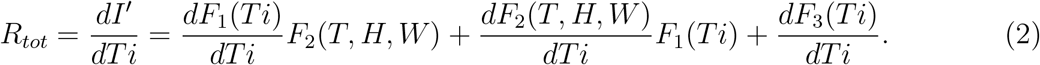

where *T, H* and *W* are assumed to vary smoothly with time *Ti*. However, under the assumption that *T, H* and *W* are not changing with time *Ti* (which is in general not true, but may be considered as an approximation for a few days when the values of *T, H* and *W* are not changing significantly) we can write *R* as partial derivative for which *R* = *R_part_* is written as

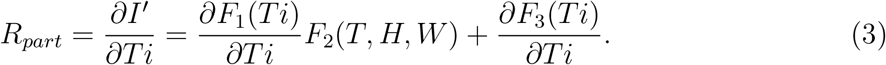

One may note that in our discussion, 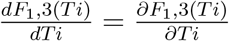. We will refer *R* as *R_part_* in general. On the right-hand side of the above equations all terms are related to the properties of the spreading of virus. To identify these features from our statistical analysis, data on the number of infection cases of different cities were taken by considering *I*′ = *I/N* corresponding to different *T, H, W, Ti* values. From the proper fitting with data points, we will determine the three functions *F*_1_, *F*_2_ and *F*_3_.

To smoothen the local fluctuations in the data (like the number of cases recorded after a day or the variations of temperature, relative humidity or wind velocity) over a short period, which should not appear as a global effect, all the data were averaged over a period of three days. These averaged data points were tried to fit with various possible functional forms of *F*_1_, *F*_2_ and *F*_3_. We have considered various forms for these functions including linear, bi-linear, tri-linear, non-linear and other simpler forms. In this work, three models are presented as shown in Table I: In Model A, *F*_1_ depends on *Ti*^2^ and *F*_2_ depends on both *T* and *H* and *F*_3_ is absent. *F*_2_ depends linearly in *T* and *H* with positive coefficients. Then there is bilinear term *TH* with negative coefficients. This is expected to be related to the condition showing the lack of stability of the virus under certain temperature and relative humidity subject to the strength of the linear terms in *T* and *H*. There are three parameters evaluated statistically considering best fit with the data and their *p* values are shown in Table I. In Model B, *F*_1_ depends on *Ti*^2.6^ and *F*_2_ depends on both *T* and *H* and *F*_3_ is absent. *F*_2_ depends linearly in *T* with positive coefficients but depends on relative humidity as *H*^1.5^. Then there is bilinear term *TH* with negative coefficients like Model A. There are three parameters evaluated statistically considering best fit with the data and their *p* values are shown in Table I. In Model C, *F*_1_ depends on Ti^2^ and *F*_2_ depends on *T, H* and *W* and *F*_3_ depends on Ti^4.^ *F*_2_ depends linearly in *T* with positive coefficients but quadratically on *H* and *W*. Then there is bilinear term T*H* with negative coefficients like other two models. There are five parameters evaluated statistically considering best fit with the data and their *p* values are shown in Table I.

**Table I.** Three Regression models. *Different regression models with the relationship of I′ with T, H, W and Ti. p values of the numerical coefficients are given in the same order as the order of the coefficients in the fitted equations*.

| Model | Best Fit Relationship for $I'$; $p$ values of coefficients | Adjusted $\bar{R}^2$ |
| --- | --- | --- |
| $Model(A)$ | $I' = (5.54765 \times 10^{-7} T + 6.14892 \times 10^{-8} H - 9.70146 \times 10^{-9} T H) Ti^2$<br>$p$ values: 0.000607788, $1.54061 \times 10^{-8}$, 0.0000169868, | 0.584545 |
| $Model(B)$ | $I' = (4.59159 \times 10^{-8} T - 9.46134 \times 10^{-10} T H + 1.00967 \times 10^{-9} H^{1.5}) Ti^{2.6}$<br>$p$ values: 0.0125458, 0.000387766, $4.16713 \times 10^{-11}$ | 0.629524 |
| $Model(C)$ | $I' = (2.46481 \times 10^{-7} T - 5.02565 \times 10^{-9} T H + 5.2107 \times 10^{-10} H^2 - 2.88696 \times 10^{-9} W^2 + 2.7413 \times 10^{-9} Ti^2) Ti^2$<br>$p$ values: 0.00382538, 0.000162158, $2.8341 \times 10^{-7}$,<br>0.0382248, $3.26429 \times 10^{-6}$ | 0.672678 |

Next, we discuss the validation of the regression models in fitting the data. In obtaining these three models we have taken care of: 1) Plot of observed and predicted response 2) Standardized residual plots showing the characteristics of errors in the model with respect to observed data 3) adjusted *R*^2^ value which indicates the goodness of fit, 4) the *p* value for the parameters appearing as coefficients with the various variables in *F*_1_, *F*_2_ and *F*_3_ which indicates what is the probability of the values of those parameters not to be so and 5) the predictive power of the model beyond data points. Here, by predictive power, it is meant that at some moderate temperature, *I*′ should not vanish for all *H* or *W*, as such things have not been observed. In some cases, the first three points are well-satisfied for some functional forms of *F*_1_, *F*_2_ and *F*_3_ but the point 5 is not satisfied and those cases were discarded. These three models satisfy point 5. Also, in obtaing these three models, we have imposed the condition *p<* 0.05 for all numerical coefficients as shown in Table I, for the model to be statistically significant. In subsequent discussions, based on the first four criteria, we will discuss merits and demerits of these three models. For statistical analysis, *Mathematica* (URL: https://www.wolfram.com/mathematica) has been used.

The best fit for the model will be determined by minimizing the weighted sum of squares of the deviation between the data and the fit. The weighted sum of squares is given by

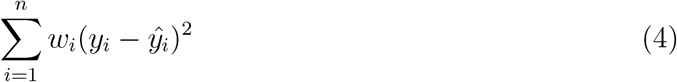

where *y_i_* is the observed and 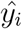 the fitted values of the response (which is *I*′ for Model A and *I*′/*Ti*^2^ for Model B and C) corresponding to the number of infected cases and *n i*s the total number of data points. It has been expected that 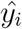 is in general function of *Ti, T, H, W* and some parameters (*a_j_* where *j* is the number of parameters) appearing as coefficients of the variables *Ti, T, H, W*. If the weights *w_i_* are same for all data points then it may be considered as 1. The hat matrix *H i*s given by 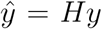 where 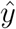 is the predicted response vector and *y* is the observed response vector. *H* is a *n × n* matrix where *n* is the number of data points. The *i*-th standardized residuals is the scaled form of the residuals and is given by (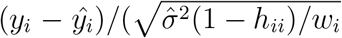 where 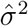 is the estimated error variance including the *i*-th data point for standardized residuals, *h_ii_* is the *i*-th diagonal element of the hat matrix *H* and *w_i_* is the weight of the *i*-th data element. To check the validity of the models, we will consider standardized residuals plots for different models as shown in Figure 1. Residuals give a measure of the pointwise difference (*y_i_* − 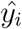) between the observed *y_i_* and the fitted values 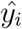 of the response corresponding to the number of infected cases. These are not true errors but estimated errors. For statistical analysis, two criterias for errors are (1) they are normally distributed and in that case, standardized residuals are preferably within ±2 in 95% cases (2) They are uncorrelated, which is expected to reflect in the standardized residual plots. These may not be totally independent for some *F*_1_ which is function of *Ti*. In that case, heteroscedastic feature will be seen in the standardized residuals. To make that somewhat homoscedastic, the consideration of some weights *w_i_* = 1 could be useful. As for example, with the type of function considered for *I*′ for model A (as shown in Table I) without different weights for different data points, the standardized residuals are found to be highly heteroscedastic. To avoid that (as required by the criterias for errors mentioned above), for model A, 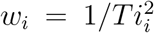 have been considered. However, for model B and C, *w_i_* = 1. In Table I, corresponding to each model, the Adjusted 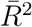 is shown which represents the goodness of fit with data and will be considered for comparing models. 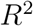 is the ratio of the model sum of squares to the total sum of squares. The Adjusted 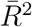 is given by 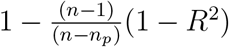 where *n_p_* is the number of parameters and with increase in *n_p_*, Adjusted 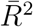 is reduced.

**FIG. 1.**
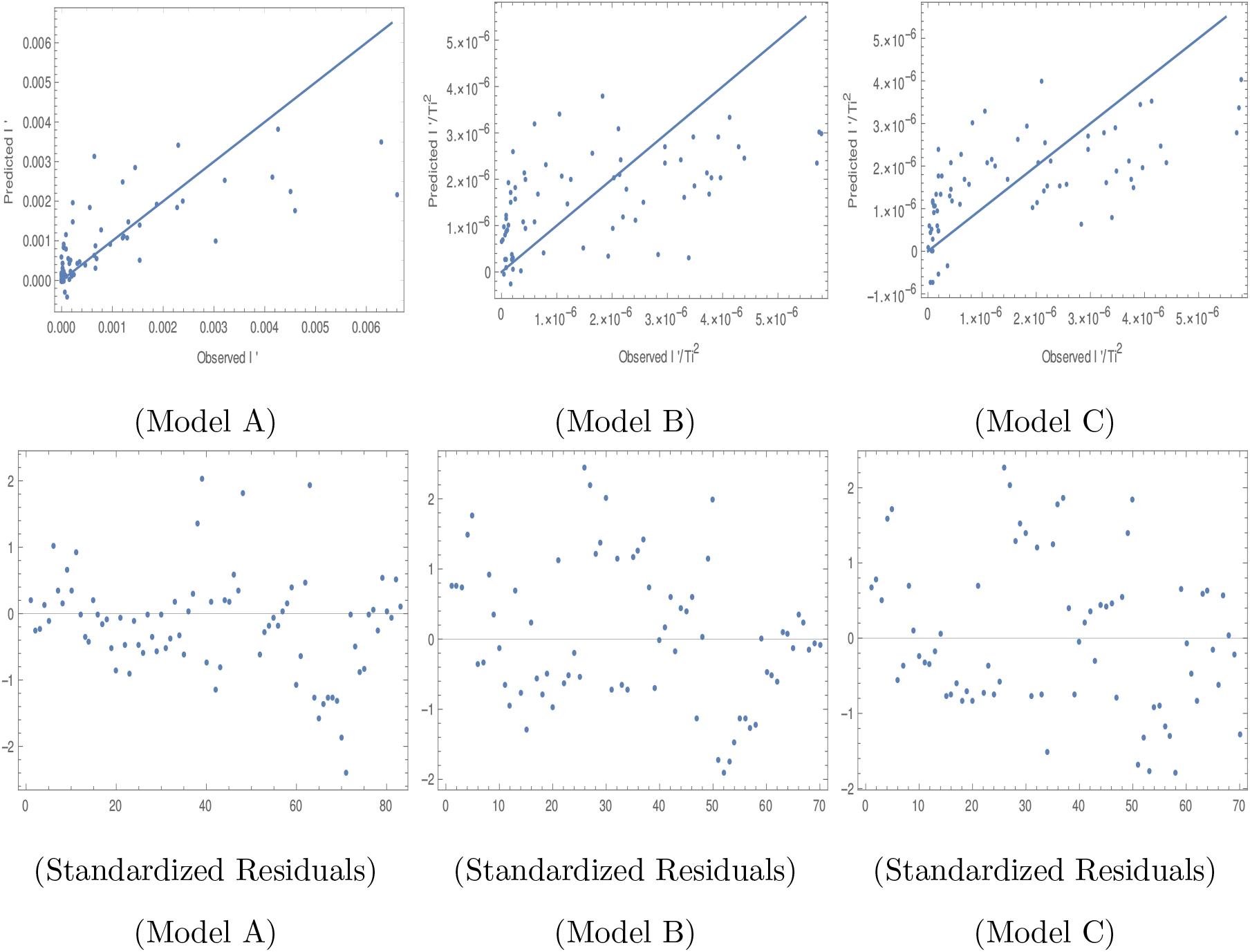
Observed versus predicted values of I′ for Model A and of *I*′ /*Ti*^2^ for Model B and C are shown in upper part of Figure. The Standardized residual plots for three models are shown in the lower part of the Figure.

In the upper part of Figure 1 the predicted versus observed *I*′ is shown for Model A. The points which are nearer to the straight line are better fit as the points on the straight line corresponds to the equality of observed and predicted values. For higher values of *I*′ the fit is not so good. For Model A, if no different weights are considered, then there is too much heteroscedastic feature in the corresponding standardized plot. Although after considering weights 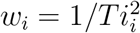 for different data, heteroscedasticity is reduced but still little bit it is there as seen in standardized residual plot for Model A. Removing some outliers or changing the functional form, does not help much when we consider also the *p* values for the parameters to be at least lesser than 0.05 and adjusted 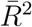 greater than 0.50. Considering various possible functional forms of F 1, *F*_2_ and *F*_3_, it is found that there should be at least overall a factor of *Ti*^2^ in fitting *I*′. Also it could be that because of this factor, the error in *F*_2_ and *F*_3_*/F*_1_ is getting magnified while considering the error in *I*′. So instead of *I*′, best fit for *I*′ /*Ti*^2^ has been tried in case of Model B and C and then better homoscedastic standardized residual plots have been found. In that sense, Model B and C are better than model A. However, we have to remove a few outliers also to achieve this homoscedastic feature. The observed versus predicted *I*′ /*Ti*^2^ plots in Figure 1 for Model B and C are better keeping in mind that the scales are different from that shown for Model A. The adjusted 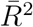 is better for Model C in comparison to other two models as shown in Table I. Combining all these points, model C seems to be better than other two models. However, it is to be kept in mind that more data points have been considered for model A. So we will discuss all these models further.

In Figure 2, the mean prediction band for *I*′ has been shown for different models for different time at 90% confidence level. However, other variables such as *T, H* and *W* have been fixed at *T* = 30°C, *H* = 40% and *W* = 10 Km/hr. For higher number of days, the band width is shortest for Model C and then comes Model A and then B so far widths are concerned. Model A and B are independent of *W*. For Model C, even if *W* is considered 40 Km/hr instead of 10 Km/hr, the band width for Model C is found to be smaller in comparison to Model A and B. This is found to be true even at 99% confidence level and also with the variation of *T* and *H* values. So Model C is also better in the sense that it has lesser uncertainty in the prediction of *I*′ as long as *W* is not above about 40 Km/hr.

**FIG. 2.**
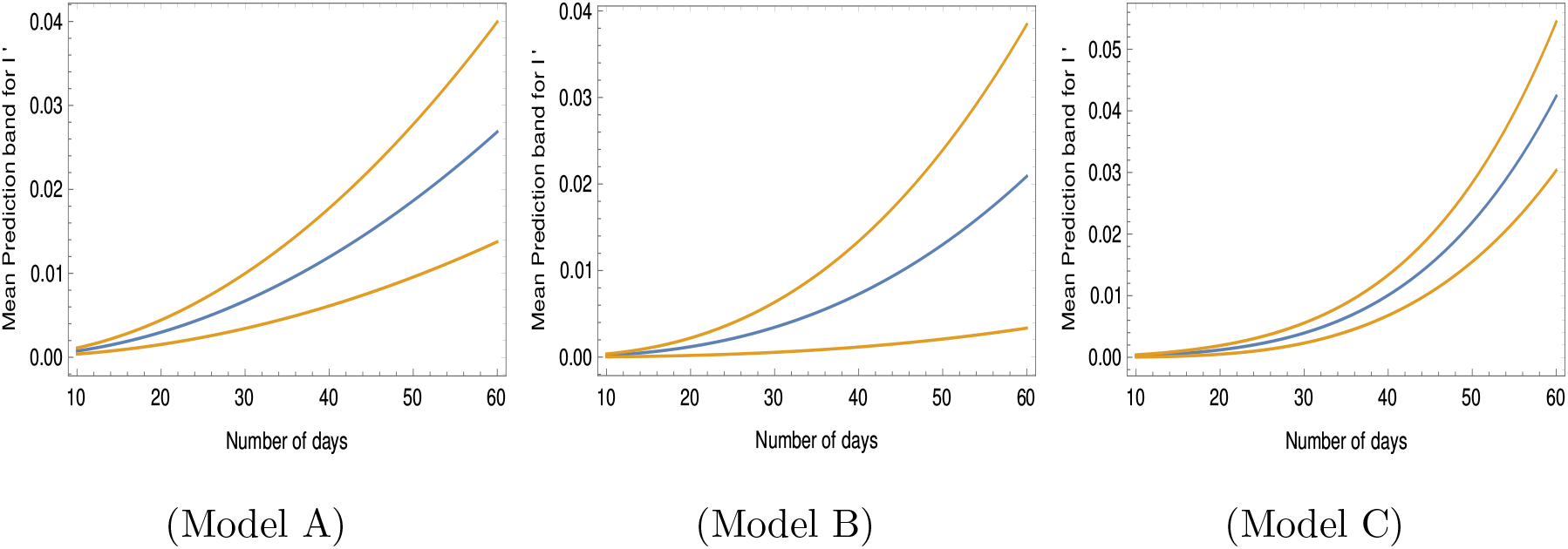
Mean prediction band for *I*′ at 90% confidence level for different models at *T* = 30°C, *H* = 40% and *W* = 10 Km/hr. Blue line corresponds to best fit values.

In all these models *I*′ varies as *Ti*^2^ or with higher powers of *Ti* and 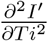 is always non-zero. For that, the rate of increase in the number of COVID-19 cases increases with at some rate. This will be further explained at the end of the result in section 4. For *F*_1_, the exponential 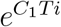 form *e*^*C*_1_*T*_i_^ (where *C*_1_ is a parameter) has also been tried. In such case, one could write 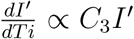 which is somewhat like SIR models considered extensively. But that kind of *F*_1_ does not give proper satistical validity of the model so far data is concerned. So it seems difficult to get SIR kind of models for the transmission of COVID-19.

In this study on transmission of COVID-19, we are concerned with the number of infected individuals. At present, there is no evidence of gendered impact on this number of infected individuals, although there seem to be some impact on the number of mortality due to the disease (Clare Wenham, Julia Smith, Rosemary Morgan, on behalf of the Gender and COVID-19 Working Group†, March 6, 2020 https://doi.org/10.1016/S0140-6736(20)305262). So our analysis is not expected to have gendered impact. The distribution of population in different age groups in Spain, Italy and the USA do not differ much. Based on World Bank Data in 2017, the percentage of population in age group of 0-14, 15-64 and 65+ in years may be considered about 15%, 65% and 20% respectively for these three countries. For Brazil, these are are about 21%, 70% and 9%. If the number of infected individuals in different age groups follow the same distribution then one may conclude that the age of individuals is not playing any role in the infection due to SARS-CoV-2 virus. However, it seems that for the age group 0-14 years the number of infected cases is much lower than 15% for all these three countries. So it could be expected that there is different infection rate in different age groups in different countries, But due to lack of availability of data of number of infected persons based on age groups, in different cities, we have refrained from doing the analysis based on age groups. We have not analysed the data of infected cases where lock-down/travel restrictions have been imposed quite early like the case of India. Depending on the nature of restrictions, probably *I*′ (as shown by the models in Table -I) is to be scaled in such cases by some overall factor.

## 4. Results for different models

The number of infected cases for a population of 5 × 10^6^ is shown in contour plots in Figure 3, 4 and 5 at different temperatures and relative humidities corresponding to model A, B and C respectively. One may compare different zones of temperature and relative humidity where the virus is expected to cause more or less infection or no infection. To get the number of cases, *I*′ has been multiplied by the total population *N*. In Figure 3, 4 and 5, we have considered *I*′ in first 20 days and first 40 days after initial infection. The plots are shown considering the same temperature and relative humidity for the entire period. We have discussed the variations of temperature and humidity in Section 5. Due to the variations in the environment in different places, the number of cases will be different than what has been shown. However, these figures will give the understanding of how virus is viable under different weather conditions.

**FIG. 3.**
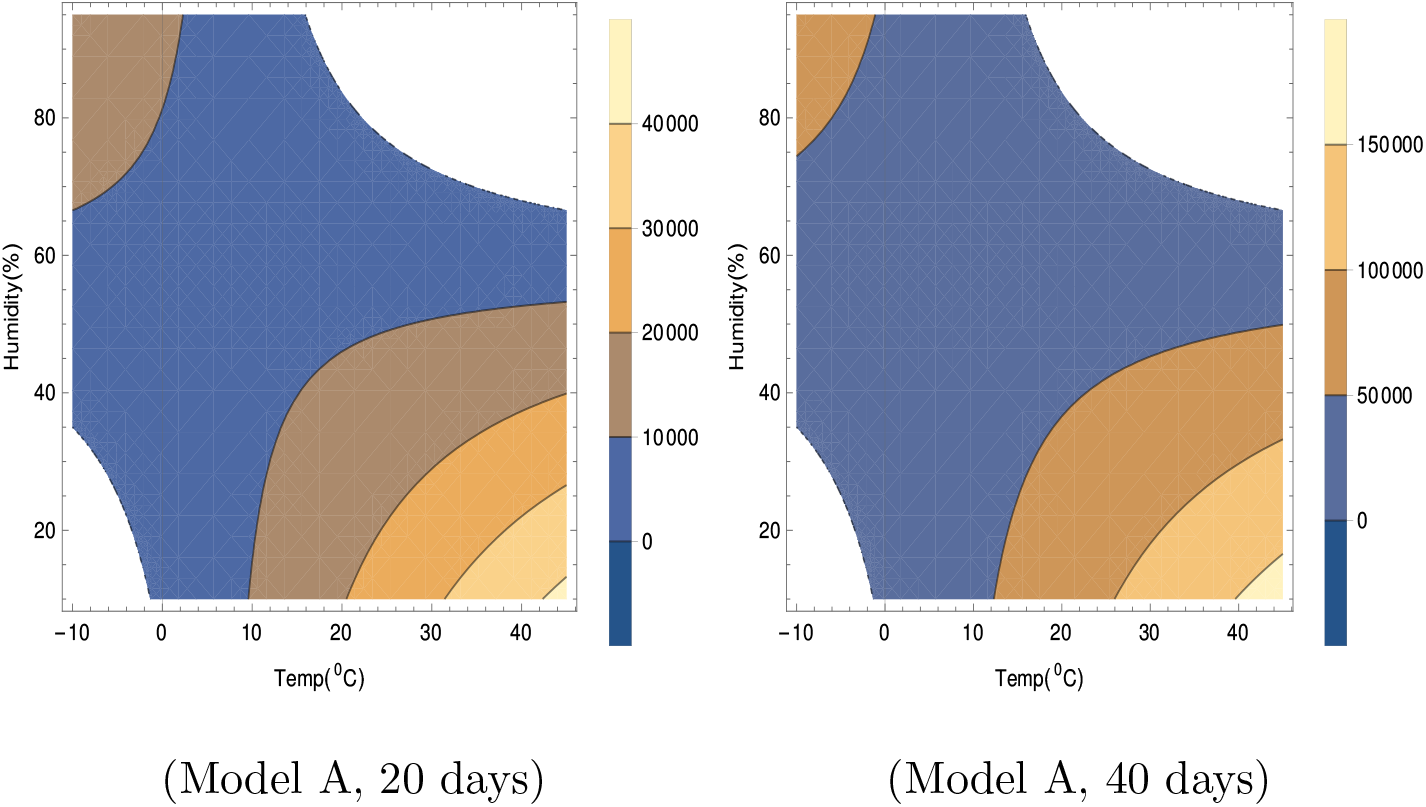
Contour plots of number of virus-infected persons with temperature and relative humidity for Model A in the first 20 days and 40 days after the initial cases of infection. Total population is 5 × 10^6^. Different colors in the legend shows ranges of the number of infected cases.

**FIG. 4.**
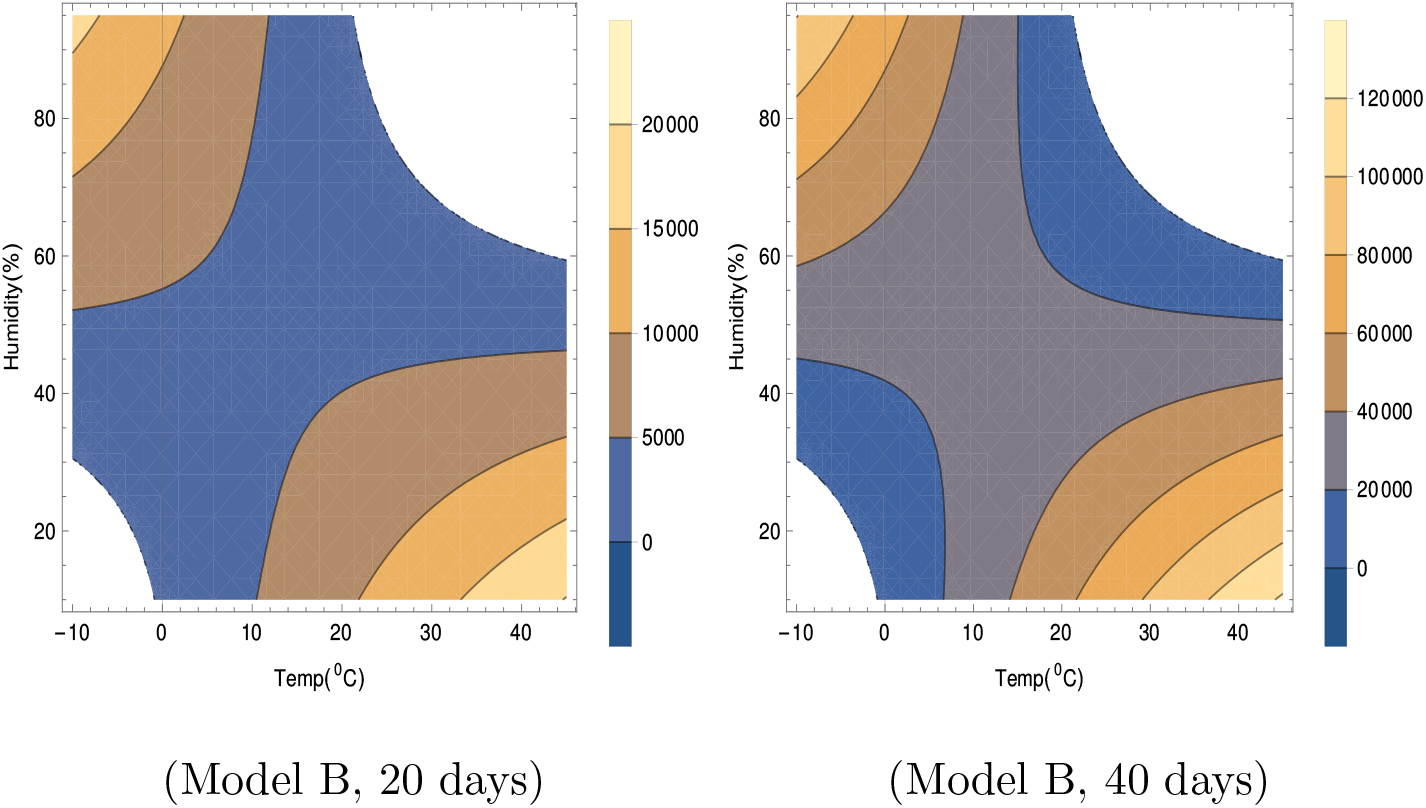
Contour plots of number of virus-infected persons with temperature and relative humidity for Model B in the first 20 days and 40 days after the initial cases of infection. Total population is 5 × 10^6^. Different colors in the legend shows ranges of the number of infected cases.

**FIG. 5.**
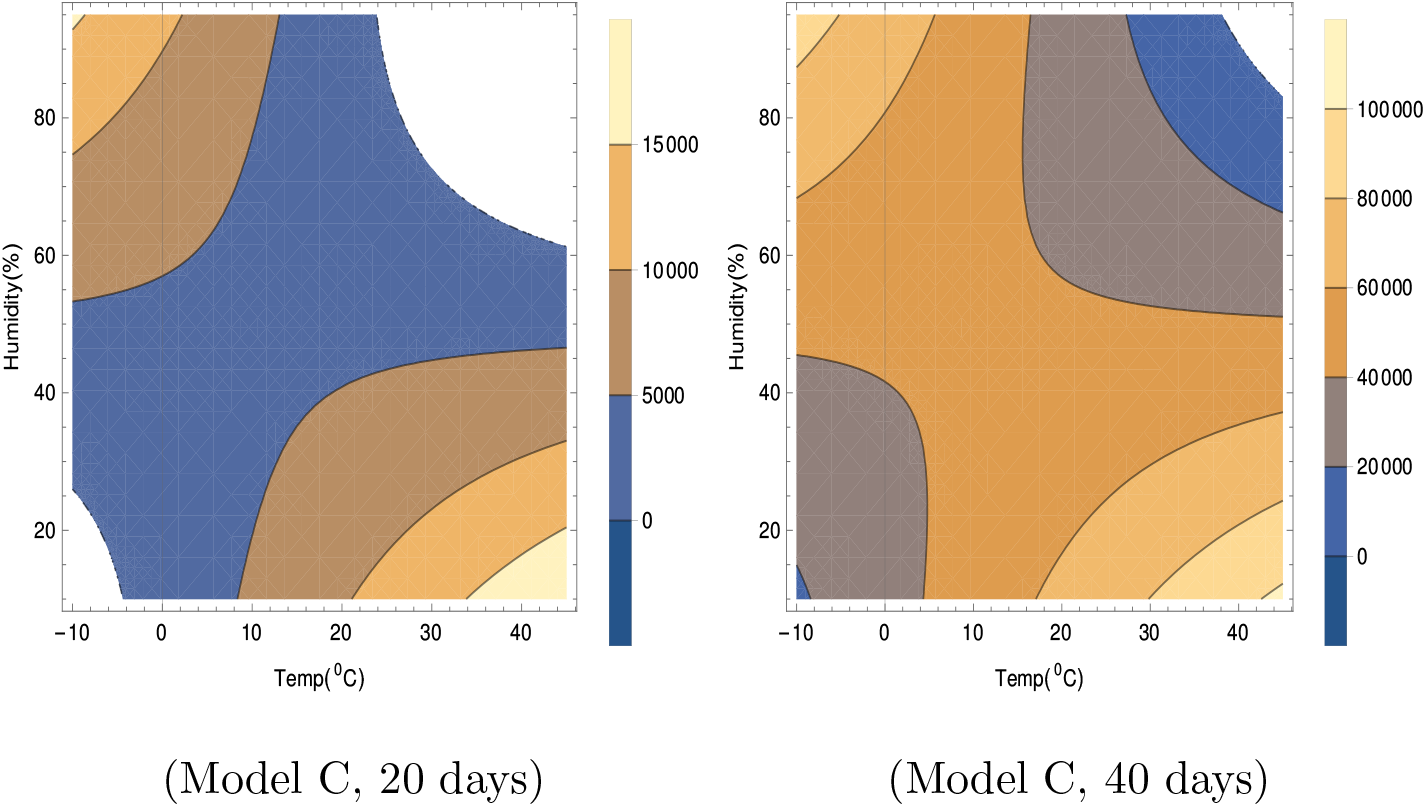
Contour plots of number of virus-infected persons with temperature and relative humidity for Model C in the first 20 days and 40 days after the initial cases of infection. Total population is 5 × 10^6^. The wind speed *w* = 10 Km/hr has been considered. Different colors in the legend shows ranges of the number of infected cases.

In Table 1, it is seen that *F*_2_ in Model A depends on *T* and *H only*. Wind speed *W doe*s not play any role. There is no *F*_3_ part in *I*′. In Model A, in Figure 3, for lower temperature around −10°C, viral infections are expected for relative humidity above about 35 %. The unshaded regions in Figure 1 correspond to absence of viral infections for the corresponding temperatures and relative humidities. The infections are not expected in the unshaded region in Figure 3, for temperature from −2^0^*C t*o −10^0^*C* and relative humidity 0 to 35%. For higher temperature, the viral infections are not expected in the unshaded region for temperature from 16^0^*C t*o 45^0^*C* and relative humidity 65 to 95%. Higher infections are expected for higher temperature and lower humidity corresponding to right lower side of both plots in Figure 3 and lower temperature and higher humidity corresponding to left upper side of both the plots. These features are true even with the increase in number of days in this model.

In Table 1, it is seen that *F*_2_ in Model B depends on *T* and *H* only. Wind speed *W* does not play any role like Model A. There is no *F*_3_ part in *I*′. However, the total number of cases as shown in Figure 4, is relatively lesser than that in Model A. In Model B, in Figure 4, for lower temperature around −10^0^*C*, viral infections are expected for relative humidity above about 30 %. The viral infections are not expected in the unshaded region in Figure 4, for temperature from −2^0^*C* to −10^0^*C* and relative humidity 0 to 30%. For higher temperature, the viral infections are not expected in the unshaded region for temperature from 22^0^*C* to 45^0^*C* and relative humidity 60 to 95%. Similar to model A, in model B also, higher infections are expected for higher temperature and lower humidity corresponding to right lower side of both plots in Figure 4 and lower temperature and higher humidity corresponding to left upper side of both the plots. These features are true even with the increase in number of days in this model.

In Table 1, it is seen that unlike earlier two models, *F*_2_ in Model C depends on *T, H* and also wind speed *W* and *I*′ in Model C, depends on *F*_3_ also. In both the contour plots in Figure 5, wind speed is fixed at *W* = 10 Km/hr. The total number of cases as shown in Figure 5, is relatively lesser than those in other two models. In Model C, in Figure 5 for the left hand side plot for 20 days, for lower temperature around −10^0^*C*, viral infections are expected for relative humidity above about 25 %. Infections are not expected in the unshaded region in Figure 5 for the left hand side plot for 20 days, for temperature from −4^0^*C* to −10^0^*C* and relative humidity 0 to 25%. For higher temperature, infections are not expected in the unshaded region for temperature from 24^0^*C* to 45^0^*C* and relative humidity 60 to 95%. Similar to model A and B, in model C also, higher infected cases are expected for higher temperature and lower humidity corresponding to right lower side of left plot in Figure 5 and lower temperature and higher humidity corresponding to left upper side of the left plot for the first 20 days. But unlike other two models, these features change with time, as can be seen from the right hand side plot in Figure 5. This is due to the presence of purely time dependent part *F*_3_ in case of Model C. Gradually the unshaded regions become smaller with increase in time and later on do not exist. So viral infections are possible for wider range of humidity and temperature in Model C with the increase of time after initial infection.

For Model C, the effect due to the variation of wind velocity in the range 0 to 25 Km/hr is shown in Figure 6. The regions for no viability of viral infections, related to lower temperature and lower relative humidity and also related to higher temperature and higher relative humidity; becomes larger for higher wind velocity. This can be seen in both left and right plots corresponding to 20 and 40 days respectively. Probably it happens because virus are blown away more by the stronger winds even to the areas where hosts do not come into contact with the virus easily. However, the total unshaded region decreases with time. So viral infections are possible for wider range of humidity and temperature as well as wind velocity in Model C with the increase of time after initial infection.

**FIG. 6.**
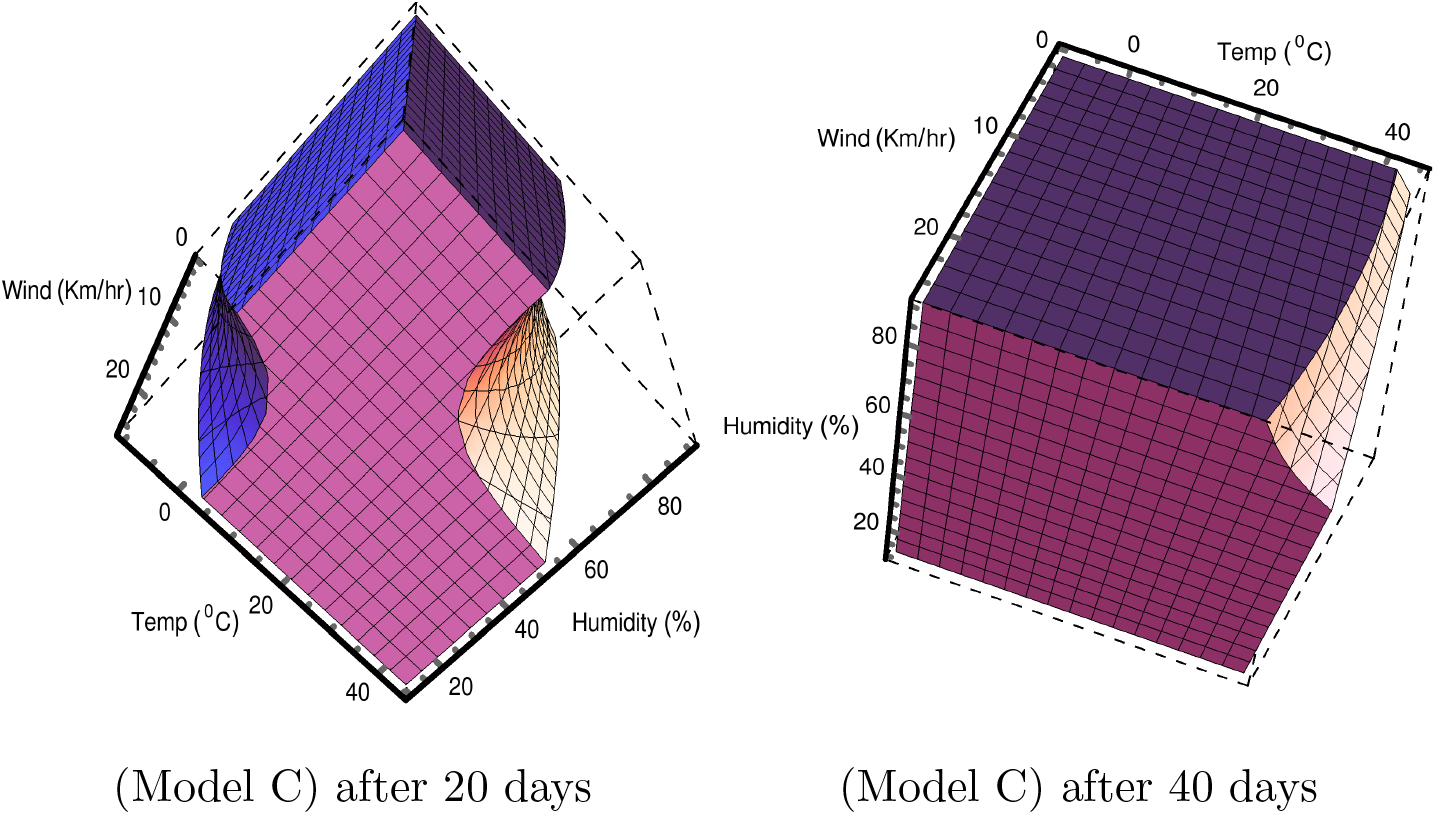
Allowed region in 3D space of temperature, relative humidity and wind velocity in different models for which number of virus infected persons are greater than 100 in first 20 days after the initial spreading. Total population is 5 × 10^6^.

The rate of increase of number of infected cases is equal to rate of increase of *I*′ times *N*. The rate of increase of *I*′ for different models can be written as

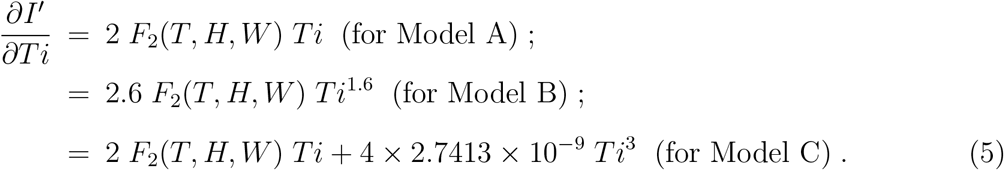

*F*_2_ in above equation are different for different models as follows from the relationship for *I′* shown in Table I. In Figure 7, the rate of increase of *I′* written as 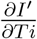 in equation (5) is shown for different models for temperature 30^0^*C* and relative humidity 50% and wind speed *W* = 10 Km/hr. One can see that for Model A, 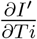 is increasing linearly with time.

**FIG. 7.**
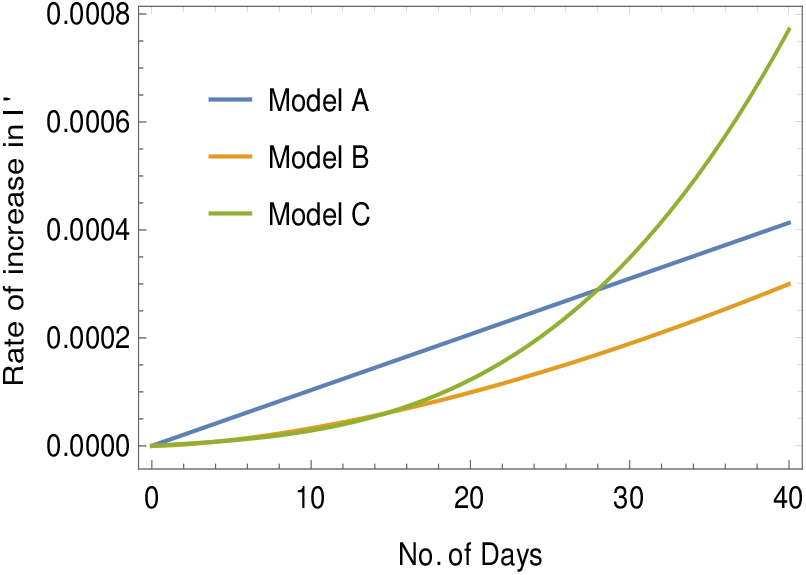
Plot of 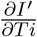 versus *Ti* for temperature 30°C, relative humidity 50% in models A, B and C. For model C, wind velocity is 10 Km/hr.

For other two models it is non-linear. But this increase is faster in Model C at a later time in comparison to other two models for the chosen temperature, relative humidity and wind speed. For Model A, the rate of increase of 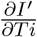 with time is 2*F*_2_. For Model B it is, 2.6 × 1.6 *F*_2_ *Ti*^0.6^ and for Model C, it is 2*F*_2_ + 12 × 2.7413 × 10^−9^ *Ti*^2.^ So for particular *T* and *H* values, this is constant in Model A. However, in Model B and in Model C, it is also varying with time as can be seen in Figure 7.

## 5. Statistical model-inspired rise and fall in the spreading of COVID-19

In Figure 8, considering model (B), an illustrative example of rise and fall of *I′* which also means the rise and fall of the spreading of the virus, has been presented. Here, the variation of temperature and relative humidity with time *Ti* has been considered. For simplicity, we have assumed the changes in temperature and relative humidity are almost instantaneous. In this example, in the first 20 days, *T* = 20^0^ C and *H* = 50 % and in the next 10 days it is *T* = 30^0^ C and *H* = 40 % and after 30 days it is *T* = 35^0^ C and *H* = 85 %. To plot *I*′ in these three time periods the following functions for *I*′ has been used:

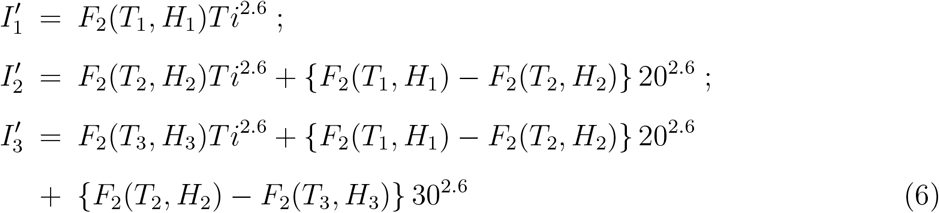

in which 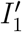 is to be considered with *Ti* varying from 0 to 20, 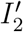 is to be considered with *Ti* varying from 20 to 30 and 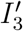 is to be considered with *Ti* varying from 30 onwards.

**FIG. 8.**
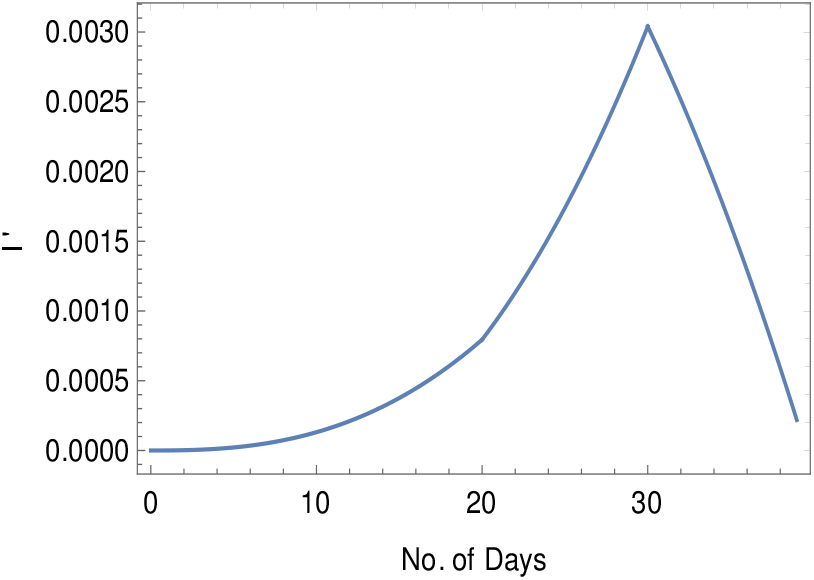
Variations in spreading of SARS-CoV-2 in terms of the variations of *I*′ according to Model B due to *T* = 20^0^ C and *H* = 50 % in first 20 days; *T* = 30^0^ C and *H* = 40 % in the next 10 days and *T* = 35^0^ C and *H* = 85 % after 30 days.

From Figure 4 for Model (B), one can see that for (*T*_2_*, H*_2_), the number of infected cases is more than that for (*T*_1_*, H*_1_). So in Figure 8, there is steeper rise in *I*′ during 20 to 30 days than that during 0 to 20 days. The (*T*_3_*, H*_3_) is in the region where viral infections are not viable in Figure 4. *F*_2_ is actually negative in this region which should be interpreted as the fall in the spreading of the virus with which *I*′ is related. Once the weather is in such no viable (unshaded) region for the SARS-CoV-2 viral infections for some days, one might expect that COVID-19 could go away. In Figure 8, the period after 30 days, corresponds to a fall in the spreading of the virus. We have considered that the function *F*_2_ (which indicates the interactions of virus with the environment), will remain the same during this period. Figure 8 is an illustrative example of how large cities could be partially free from COVID-19 in a natural way due to the change in the meteorological factors in the environment. This is possible in Model A also. But in Model C this can happen only in very early stage but later on it is not expected as the unshaded region will not be found later as shown in right plot in Figure 5. The possiblity of a fall in COVID-19 cases has been discussed due to only meteorological factors. But this could also be due to the development of immunity to SARS-CoV-2 in the human body. No analysis on that aspect has been done here.

## 6. Connection of transmission characteristics with the structural components of the SARS-CoV-2 virus

The statistical analysis through function *F*_2_ indicates a complex interaction between COVID-19 infection and meteorological parameters. The infection is expected to enhance under two different combinations of environmental conditions i.e. lower temperature-higher humidity and higher temperature-low to ambient humidity as found in Figures 3-5. Such nature of interaction can be explained by the physicochemical characteristics of respiratory droplets or aerosol and the structural features of the virus itself. The enveloped virus SARS-CoV-2 has a positive sense, single-stranded RNA genome which encode four important structural proteins, namely spike glycoprotein (S), envelope protein (E), matrix glycoprotein (M), and nucleocapsid protein (N). The S glycoprotein is surface exposed and mediates the viral entry into host cells. It comprises two regions S1 and S2, where S1 is responsible for binding to the receptor of the host cell and S2 is for the fusion of the viral and cellular membrane. Although both SARS-CoV, SARS-CoV-2 attach the host cells through the binding of receptor-binding domain (RBD) of S1 region to the angiotensin converting enzyme 2 (ACE2) (Schoeman and Fielding, 2019; Walls et al, 2020), the pandemic impact of the later happens to be more severe with continuous worldwide increase in the number of infection cases and mortality.

Several factors including, the structural features of virus for persistence in the environment and for human contact might be associated with the severe infection of SARS-CoV-2. Recently, an experimental study based on Cryo-EM structure of the SARS-CoV-2 spike protein in prefusion conformation has suggested higher binding affinity of SARS-COV-2 with human ACE2 than SARS-COV (Wrapp et al., 2020). Another study on the protein-protein interaction and molecular dynamics simulations of RBD-ACE2 complex for SARSCoV-2 and SARS-CoV showed significantly lower binding free energy of the SARS-CoV-2 RBD-ACE2 interaction (−50.43 kcal/mol) compared with SARS-CoVRBD-ACE2 interaction (−36.75 kcal/mol) and thus suggesting higher binding affinity of SARS-CoV-2RBD-ACE2 interaction (He et al., 2020). Ou et al., (2020) analyzed the SARS-CoV-2RBD mutations worldwide and found the equilibrium dissociation constant of three RBD mutants to be two orders magnitude lower than the prototype Wuhan-Hu-1 strain indicating remarkable increase in the infectivity of the mutated viruses.

Apart from the increased binding affinity of SARS-CoV-2 to ACE2, the severity of COVID-19 can also be determined by the stability of the virus in the environment. Although most of the studies on the effect of meteorological factors on the COVID-19 have speculated the decline in infection with an increase in environmental temperature and humidity (He et al., 2020; Wang et al., 2020), the number of infection cases in reality is increasing sharply with the rise in temperature. Present study has also indicated such increase at higher temperature as the general feature of the models (Table I) particularly for lower humidity. The persistence of the virus at higher temperatures can be explained in the context of the stability of the spike protein because it is a critical component determining the infectivity. A lengthy molecular dynamics simulation of trimeric spike proteins of SARS-CoV-2 and SARS-CoV has shown that the spike protein of SARS-CoV-2 has significantly lower total free energy (−67,303.28 kcal/mol) than the spike protein of SAR-CoV (−63,139.96 kcal/mol). Similarly, the free energy of the RBD of SARS-CoV-2 spike protein is relatively lower than that of SARS-CoV. The results thus explained increased stability of SARS-CoV-2 spike protein at higher temperature (He et al. 2020). Several studies have been carried out on the effect of relative humidity on the survival and infectivity of enveloped and non-enveloped viruses (Benbough 1971; Shaman and Kohn; Pica and Bouvier, 2012; Yang and Marr, 2012; Marr et al. 2019). When the virus is released into the environment as part of a respiratory fluid droplet, relative humidity of environment controls the amount of water evaporated from the droplet until equilibrium with the surrounding air is maintained/reached (Prussin et al., 2018). The respiratory viral droplets or aerosol on exposure to lower to ambient relative humidity environment are subject to evaporation due to the vapour pressure gradient between its surface and air. As evaporation proceeds, the water vapour pressure at droplet surface decreases. The presence of lipid membrane in the enveloped viruses has been evidenced to protect their capsids from damage due to change in humidity leading to survival at lower humidity conditions (Benbough, 1971; Yang and Marr, 2012). Hence, a considerable higher infection level of SARS-CoV-2 at low to ambient relative humidity and higher temperature as predicted in the present study could be related to the presence of structural features of the viral envelope and relatively stable spike protein with significantly lower free energy. Moreover, at lower relative humidity and higher temperature the respiratory droplet will undergo evaporation at a higher rate and the desiccated state of droplet will remain unaffected by the changes in temperature, causing higher infection rates. Infection can be further enhanced due to higher mobility of the smaller droplets under high temperature and low humidity which can readily enter the human host. As predicted in this study, the experiments conducted by Prussin et al. (2018) showed that at 37^0^*C temperatur*e Phi6, a surrogate of influenza and coronaviruses, had the highest infectivity at 20-40% relative humidity. This happens in all the models as shown in Figures 3-5. However, in the same experiment the virus showed a significantly higher level of infection throughout the entire range of relative humidity at lower temperature of 14 and 19°C. Particularly for Model B and C, this happens for lower temperatures 10 to 20^0^*C throughou*t the entire range of humidity as seen in the left plots in Figure 4 and 5 in the first 20 days although with some variation in the level of infection in comparison to that found for surrogate of influenza and coronavirus.

On the other hand, our statistical analysis also showed an increase in viral infection at low temperature and high relative humidity as observed earlier for seasonality of influenza virus infection, which occurs at a significantly higher level during winters when average outdoors daily temperatures remains lower and relative humidity is higher (Lowen and Steel, 2014). The finding is also supported by the results of Prussin et al. (2018) reporting higher infectivity of Phi6 at lower temperature (10−20^0^*C*) when relative humidity was kept constant at 75%. However, according to models in this work, for relative humidity at 75%, increase in viral infection occurs at further lower temperature than that found for influenza virus. It is below 0°C in Model A, and in Model B and C, below 10°C as seen in Figures 3-5 in first 20 days. Probably at higher humidity with appropriate droplet size SARS-CoV-2 could remain wet by keeping it at slightly higher temperature with respect to the lower surface temperature of the droplet and thus remains viable for infection.

However, our study indicates a complete reduction in infectivity of the virus under lower temperature and lower humidity condition. Under such condition, probably SARS-CoV-2 virus could not remain wet inside the droplets which are of smaller size and the lipid and the protein structure inside the virus gets deformed resulting in a reduction in viability.

Furthermore, the predicted decrease in viral infectivity at a higher temperature and higher humidity in our models could be due to the reduced rate of evaporation of respiratory droplets at higher humidity, which consequently makes them highly susceptible to higher temperature resulting in non-viability of SARS-CoV-2 for infection. Furthermore, corresponding aerosols with bigger size falls down to the surface and loses the mobility. This also could be a reason for the reduction in the viability of infection of SARS-CoV-2 at both high temperature and high humidity as indicated by the models.

## 7. Conclusion

Based on statistical significance, three models may be considered to understand the future outcome of COVID-19 throughout the world and among them Model C seems better. We have not considered the data of all COVID-19 affected countries. However, based on Figure 1, it seems that data of different cities of four countries, can be considered in a single framework of a statistically validated regression model for the evolution of *I*′. Furthermore, as *I*′ is normalized to 1 for any place, it is expected that such models would give a reasonably good estimate for the evolution of *I*′ for the major cities of other countries.

Based on the general features of all the models, it appears that higher precautionary measures would be required in the cities during the summer season with higher temperature and lower relative humidity, and during winter with low temperature and high relative humidity. On the other hand, the viability of SARS-CoV-2 seems to be reduced at higher temperatures with higher humidity and at a lower temperature with lower humidity conditions in the environment, as depicted by the unshaded regions of Figures 3-5. However, Model C indicates this feature to be valid in the early infections only and may not continue for prolonged period.

Based on models there is some non-zero rate of increase of 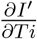 with time, which indicates some acceleration in the spread of the virus. This is related to the function *F*_2_ in Model A and functions *F*_1_, *F*_2_ in Model B and *F*_2_ and *F*_3_ in Model C. These features are expected to be valid for the spreading of other types of viruses which infect the human host. However, further studies on exploring such acceleration and the relationship of *F*_2_ with the structural features of SARS-CoV-2 would be worthwhile.

## Data Availability

All data used for analysis were available in public databases. The data on the number of COVID-19 infection cases of several major cities in Spain, Italy,the USA and Brazil (Countries with relatively severe COVID-19 outbreak) from the following sources - (1) Data of Spain (URL: https://www.mscbs.gob.es/profesionales/saludPublica/ccayes/alertasActual/nCov-China/documentos/Actualizacion_122_COVID-19.pdf) were collected from the database of the Centro de Coordinac\'i on de Alertas y Emergencias Sanitarias (CCAES), Spain. The center is responsible for coordinating information management and supporting the response to national or international health alert or emergencies, (2) Data of Italy (URL: http://www.salute.gov.it/portale/nuovocoronavirus/dettaglioContenutiNuovoCoronavirus.jsp?area=nuovoCoronavirus&id=5351&lingua=italiano&menu=vuoto) were collected from the database of The Ministry of Health (Italian: Ministero della Salute), which is a governmental agency of Italy and is led by the Italian Minister of Health. (3) Data on different cities of USA (URL: https://usafacts.org/visualizations/coronavirus-covid-19-spread-map/) were collected from USAFacts, which is a not-for-profit, nonpartisan civic initiative providing the most comprehensive and understandable source of government data available in the US. Data on cumulative COVID-19 positive cases, population, and population density for Sau Paulo, Rio de Janeiro, and Brasilia, Brazil, were collected from Wikipedia (https://en.wikipedia.org/wiki/COVID-19_pandemic_in_Brazil/Statistics). Meteorological data for all above-mentioned locations were collected from the World Weather Online database (URL: www.worldweatheronline.com).

https://www.mscbs.gob.es/profesionales/saludPublica/ccayes/alertasActual/nCov-China/documentos/Actualizacion_122_COVID-19.pdf

http://www.salute.gov.it/portale/nuovocoronavirus/dettaglioContenutiNuovoCoronavirus.jsp?area=nuovoCoronavirus&id=5351&lingua=italiano&menu=vuoto

https://usafacts.org/visualizations/coronavirus-covid-19-spread-map/

https://en.wikipedia.org/wiki/COVID-19_pandemic_in_Brazil/Statistics

## CRediT author statement

**Atin Adhikari:** Conceptualization, Resources, Writing - Review & Editing. **Shilpi hosh:** Conceptualization, Validation, Writing - Review & Editing. **Moon M. Sen:** Writing - Review & Editing. **Rathin Adhikari**: Conceptualization, Methodology, Formal nalysis, Writing - Review & Editing.

## Acknowledgement

We are grateful to USAFacts, U.S.A., the Centro de Coordinać ı on de Alertas y Emergencias Sanitarias (CCAES), Spain, Ministero della Salute, Italy, and World Weather Online for providing access to their collected data. This research did not receive any specific grant from funding agencies in the public, commercial, or not-for-profit sectors.

